# Clinical validation of whole-genome and transcriptome sequencing for genetic diagnostics in adult and pediatric AML

**DOI:** 10.64898/2026.11.16.26367955

**Authors:** Anna Staffas, Lucia Cavelier, Kajsa Olde, Christina Orsmark-Pietras, Tatjana Pandzic, Irina Golovleva, Linda Olsson-Arvidsson, Aleksandra Krstic, Sailendra Pradhananga, Nina Hollfelder, Khurram Maqbool, Mathias Johansson, Panagiotis Baliakas, Leonie Saft, Vladimir Lazarevic, Martin Jädersten, Stefan Deneberg, Anna Robelius, Martin Höglund, Gustav Orrsjö, Jonas Abrahamsson, Cornelis Jan Pronk, Josefine Palle, Hartmut Vogt, Karin Belander Strålin, Ulrika Norén-Nyström, Lars Palmqvist, Valtteri Wirta, Gisela Barbany, Thoas Fioretos, Linda Fogelstrand, Richard Rosenquist

## Abstract

Diagnosis and risk stratification of acute myeloid leukemia (AML) rely on the detection of diverse genetic aberrations using multiple standard-of-care (SoC) assays. With the increasing number of clinically relevant alterations and limited resolution of SoC methods, whole-genome and transcriptome sequencing (WGTS) represents a promising alternative. In this multi-center study, we evaluated the technical feasibility and clinical performance of WGTS across five sites in Sweden by comparison with SoC diagnostics. A total of 235 patients (197 adults and 38 children) underwent parallel SoC testing and WGTS analysis. WGTS identified 57/58 (98%) clinically relevant gene rearrangements detected by SoC and revealed an additional 12 rearrangements, including *RUNX1*::*RUNX1T1*, *KMT2A* and *NUP98* rearrangements. For structural variants and copy-number variants, WGS showed 90% concordance with SoC, with discrepancies mainly involving subclonal events or tetraploidy. For gene mutations, WGS achieved 94% sensitivity, detecting all *NPM1, CEBPA* bZIP, and *IDH1/2* mutations, but missing low-frequency *FLT3*-ITDs. Tumor-only analysis performed comparably to paired analysis for clinically relevant variants. Diagnostic classification was highly concordant between WGTS and SoC (WHO: 94%; ICC: 93%), with improved genetic subclassification in 5–6% of cases by WGTS. ELN risk stratification was concordant in 97% with WGTS reclassifying 2% of patients. Finally, a ring trial involving seven cases demonstrated complete agreement across sites in identifying aberrations. In conclusion, WGTS is a feasible and accurate comprehensive diagnostic approach for AML in a decentralized setting, improving detection of clinically relevant aberrations. However, complementary high-sensitivity assays for *FLT3-*ITD remain necessary for routine clinical implementation.

**Key points:**

- Whole-genome and transcriptome sequencing (WGTS) uncovers all clinically relevant variants in AML detected by standard-of-care (SoC) assays, with the exception of *FLT3*-ITDs at low allele frequency, which require sensitive targeted assays.
- Compared with SoC assays, WGTS improves detection of clinically relevant gene rearrangements, resulting in a 5–6% increase in genetically based AML diagnoses.
- Nationwide multicenter validation demonstrates that WGTS can be implemented in a decentralized setting.

## Introduction

Acute myeloid leukemia (AML) is a clonal malignant disorder arising from myeloid hematopoietic stem or progenitor cells, following the acquisition of disease-driving genetic alterations. Current classification systems, including the 5th edition of the World Health Organization classification of Haematolymphoid tumors (WHO 5^th^) and the International Consensus Classification (ICC), recognize numerous genetically defined AML entities^1,2^. Recurrent genetic abnormalities, including gene fusions, single nucleotide variants (SNVs), small insertions/deletions (indels), and copy-number variants (CNVs), are central to diagnosis, prognosis, and therapeutic decision-making, as reflected in current guidelines such as those from the European LeukemiaNet (ELN)^3^. In clinical practice, detection of these aberrations requires multiple complementary methods, such as chromosome banding analysis (CBA), fluorescent in situ hybridization (FISH), polymerase chain reaction (PCR) with fragment-length analysis, reverse transcription-PCR (RT-PCR), and targeted next-generation sequencing (NGS) gene panels.

As the number of clinically relevant genetic markers continues to grow, a targeted multi-assay approach is becoming increasingly challenging in terms of reagent costs, staff, and turnaround time. Meanwhile, comprehensive sequencing techniques such as whole-genome sequencing (WGS) and whole-transcriptome sequencing (WTS) are becoming more available and less costly. We and others have recently demonstrated that WGS effectively can identify structural variants (SV), CNVs, SNVs and indels in hematologic disorders^4–12^. Similarly, WTS can enable detection of fusion genes, differential gene expression, and aberrant transcripts^9,11–13^. Together, WGS and WTS provide an attractive alternative to the current multi-assay diagnostic workflow, possibly with higher diagnostic accuracy and the advantage of enabling discovery of new variants with potential clinical impact.

Here, we report results from a national, prospective multi-center study of acute leukemias conducted by Genomic Medicine Sweden (GMS), in close collaboration with the Clinical Genomics Platform at Science for Life Laboratory (SciLifeLab)^14,15^. The aim was to compare the detection of genetic alterations by combined WGS and WTS, collectively termed WGTS, with standard-of-care (SoC) genetic diagnostics, in patients with AML across the entire age spectrum, from children to adults, and to assess the impact of WGTS on diagnostic yield, according to WHO and ICC classifications, and ELN risk stratification. In contrast to previous studies conducted primarily at single centers, we also evaluated the feasibility of implementing WGTS nation-wide in a decentralized setting.

## Methods

### Patients and samples

The study included 235 patients (197 adults and 38 children) diagnosed with AML according to the WHO 5^th^ ed.^2^ and/or ICC^1^, recruited from six university hospitals in Gothenburg, Lund, Linköping, Stockholm, Umeå, and Uppsala. Of these, 198 cases (175 adults and 23 children) were enrolled in the national acute leukemia study between 2021-2024 (International Standard Randomised Control Trial. No ISRCTN66987142), 22 were adult biobank samples from patients diagnosed in Lund between 2019-2020 (WGS results from these are also reported in Stanek et al, 2026, preprint medRxiv), and 15 were pediatric biobank samples from patients diagnosed in Gothenburg between 2013-2018. Inclusion criteria required that SoC genetic diagnostics had been performed as part of routine clinical care and that DNA for WGS was available from either bone marrow aspirates (*n* = 231) or peripheral blood (*n* = 4). For 175 (74%) patients, DNA from non-tumor tissue was available for paired tumor-normal WGS analysis (cultured fibroblasts from skin biopsies (*n* = 74), magnetically sorted T cells from peripheral blood (*n* = 56), remission peripheral blood or bone marrow (*n* = 45)). For 193 (82%) patients, RNA from leukemic cells was available for WTS (Supplementary table 1). Tumor samples were collected as part of routine clinical diagnostics, whereas samples for germline analysis were collected either for study purposes only, or as part of local clinical routine. DNA and RNA were extracted according to standard clinical protocols at each participating laboratory. The study was conducted in accordance with the Declaration of Helsinki. Written informed consent was obtained from patients or when applicable, from their legal guardians. Ethical approvals were obtained from the Swedish Ethical Review Authority (2020-06673, 2021-04135) and regional ethical review boards (Gothenburg 1138-17, 313-18, Lund 2011/289).

### Standard-of-care (SoC) diagnostics

Cytogenetic and molecular analyses were performed according to standard clinical protocols at five diagnostic centers (Gothenburg, Lund, Stockholm, Umeå, and Uppsala). SoC testing included CBA, FISH, PCR-based detection of *FLT3* internal tandem duplications (*FLT3*-ITDs), and targeted NGS gene panels, Genomic Medicine Sweden Myeloid Gene Panel (GMS-MGP)^16^, Archer VARIANTPlex Myeloid Panel (Integrated DNA Technologies), or TruSight Myeloid Panel (Illumina) for remaining gene mutations. A complex karyotype was defined as ≥3 unrelated chromosome abnormalities in the absence of concurrent AML-defining gene-rearrangements.

### Whole-genome and transcriptome sequencing (WGTS)

WGTS were performed at four of the five diagnostic centers (Supplementary Table 2), while the remaining center sent material for WGTS to one of the other centers. Genomic DNA (target input 1000-1500 ng) was mechanically fragmented (Covaris) and prepared using the TruSeq DNA PCR-free Library Prep Kit (Illumina) or NxSeq AmpFREE Low DNA (Lucigen). mRNA libraries were generated from total RNA (target input 300-1000 ng) using the TruSeq Stranded mRNA or mRNA Stranded Kit (Illumina). Indexed libraries were sequenced as 2 × 150 bp paired-end runs on NovaSeq 6000 or NovaSeq X instruments (Illumina). For WGS, the target coverage depth was 90x for tumor samples and 30x for germline samples, resulting in median coverage (mean per sample) of 116x (range, 42x – 210x) and 37x (range, 24x – 64x), respectively. For WTS, the target sequencing depth was at least 50 M read pairs (r-p) resulting in a median of 81 M r-p (range, 20 – 228 M). Reads were aligned to either the GRCh37 (hg19) or GRCh38 (hg38) reference genomes, followed by standard workflows for quality control, duplicate removal, and variant calling (Supplementary Table 2). Filtered data were curated by local clinical laboratory geneticists.

### Data collection

A standardized study protocol form was used to collect SoC and WGTS data from the five diagnostic centers. A predefined set of genetic aberrations was required to be reported for each case (Supplementary Table 3). For SoC testing, gene mutations with a variant allele frequency (VAF) ≥5%, and *FLT3*-ITDs with an allelic ratio (AR) ≥0.01, were considered positive. Following data collection and central review, underlying data from SoC and/or WGTS were reinvestigated when needed, for example in cases with discrepant results.

### Diagnostic classification and risk stratification

Cases were classified according to WHO 5^th^ and ICC criteria^1,2^ with the exception of diagnostic qualifiers related to prior cytotoxic therapy, antecedent hematologic malignancy or germline predisposition, which were not considered. Risk stratification was performed according to ELN 2022^3^ and for the children, also as in the CHIP-AML22 trial^17^. For risk stratification based on *FLT3*-ITD, only mutations with AR ≥0.05 were considered. Co-occurring *CEBPA* bZIP-inframe and *FLT3*-ITD mutations were stratified as intermediate risk, and concurrent *NPM1* and *TP53* mutations (VAF >10%) as high risk.

## Results

### Characteristics of patients and samples

This study compared WGTS and SoC results from 235 patients with AML; 197 (84%) adults (≥18 years old) with a median age of 63 years at diagnosis (range, 19-90 years, 102 female/95 male), and 38 (16%) children with a median age of 6 years at diagnosis (range, 1-17 years, 16 female/22 male). The tumor samples analyzed had a blast cell proportion of 5-95%, median 51%, based on morphology (data available for 230 samples).

### Improved detection of diagnostic and risk-stratifying gene rearrangements with WGTS

To evaluate the capacity of WGTS to detect clinically relevant gene rearrangements, WGTS results were compared with those obtained by SoC CBA and/or FISH. The comparison included gene rearrangements relevant to AML classification according to the WHO 5^th^ and/or ICC, or ELN risk stratification. Among 235 patients, SoC diagnostics detected 58 such rearrangements, of which WGS identified 57 (98%) and additionally detected 12 rearrangements not detected by SoC, of which 3 were in children (Figure 1). These included four *KMT2A* rearrangements, two *NUP98* rearrangements, two *DEK*::*NUP214* and one each of *RUNX1*::*RUNX1T1*, *KAT6A*::*CREBBP, RUNX1*::*CBFA2T3*, and *PICALM*::*MLLT10*. Of the 70 cases with rearrangements identified by SoC and/or WGS, WTS could be performed on samples from 62 (89%) cases and confirmed all WGS-detected rearrangements predicted to generate fusion transcripts. As expected, WTS did not detect fusion transcripts in cases with *GATA2*::*MECOM* (*n* = 3) or t(7;12)(q36.3;p13.2) (*n* = 2). In addition, one *KMT2A* rearrangement detected in SoC by FISH was neither identified by WGS nor WTS. This sample showed extensive structural aberrations with rearrangements or genomic loss involving at least chromosomes 4, 5, 7, 11, 14, 17, 19, and 21. Notably, a breakpoint located ∼250 kb upstream of *KMT2A* may have generated a false-positive split signal by FISH, leading to a potential false positive result in SoC.

**Figure 1.**
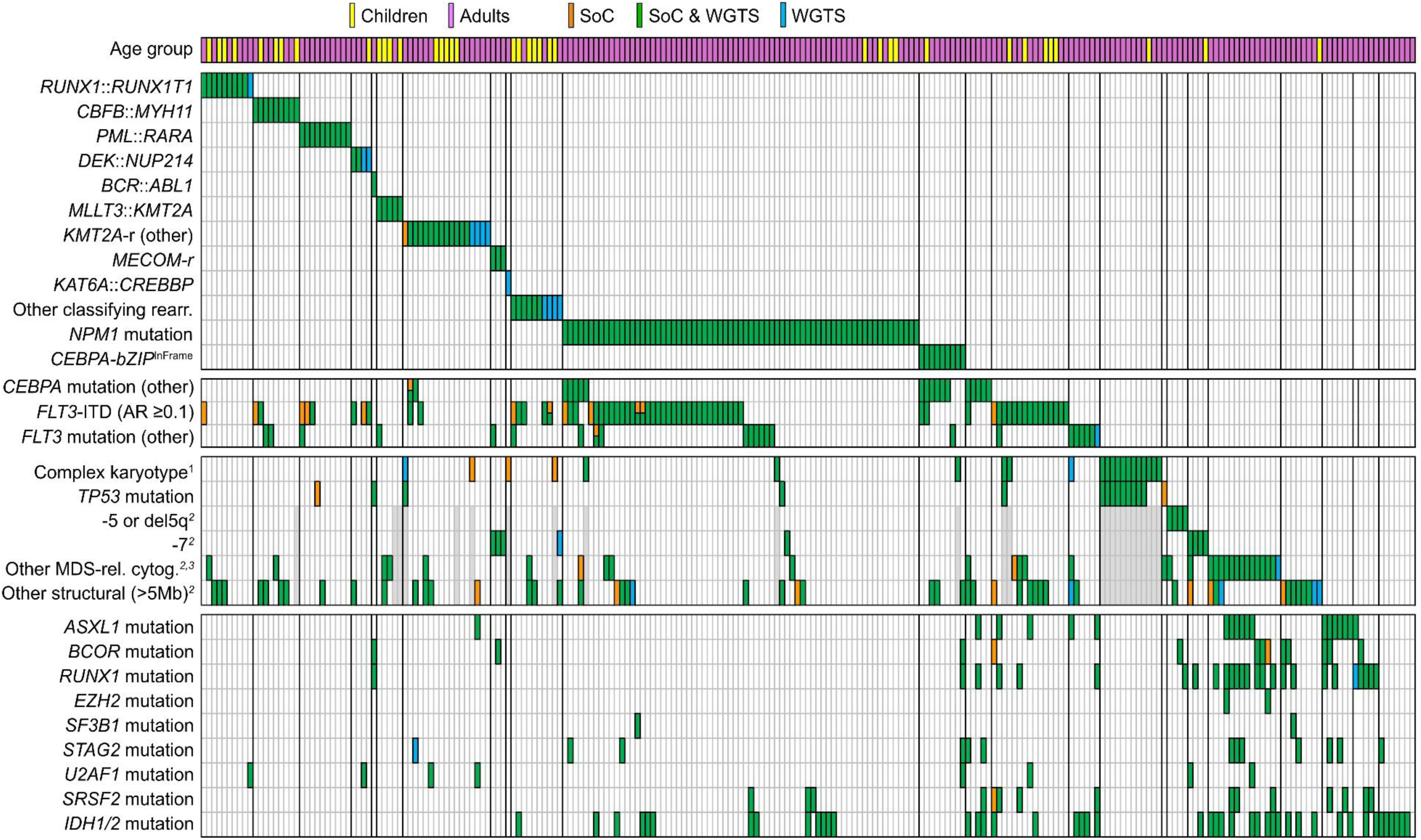
Genetic findings by SoC and by WGTS. Each column represents one AML case, total number of cases 235, children (0-17 years at diagnosis) *n* = 38 (yellow), adults (≥18 years at diagnosis) *n* = 197 (purple). Filled bars represent findings by SoC only (yellow), SoC and WGTS (green), and WGTS only (blue). ^1^Only cases without a recurrent classifying rearrangement were defined as complex. ^2^In the absence of a karyotype with ≥ 3 large CNVs or structural aberrations by SoC (such cases are marked grey). ^3^t(5q), del(7q), +8, del(12p)/t(12p)/add(12p), del(11q), −13/del(13q), del(20q) or idic(X)(q13).

In summary, WGTS improves the detection of diagnostic or risk-stratifying gene rearrangements compared with SoC.

### Other structural variants and large copy-number-variants are well detected by WGS

To assess the detection of other SVs and CNVs by WGTS, we focused on variants of importance for risk-stratification in AML and for diagnosis of myelodysplasia-related AML (AML, MR). In 20/235 patients, SoC detected CNVs and/or non-classifying SVs fulfilling the criteria for complex karyotype, including the absence of a concurrent AML-classifying gene rearrangement. In all 20 cases, WGS identified ≥3 large CNVs and/or SVs. However, in three of these cases, WGS additionally identified an AML-classifying gene rearrangement (*RUNX1*::*CBFA2T3*, *KMT2A*::*MLLT10* or *KAT6A*::*CREBBP,* respectively), resulting in reclassification. In addition, WGS identified a complex karyotype in two cases not identified as such by SoC. In the first case, described above, WGTS did not support a *KMT2A* rearrangement but instead showed aberrations consistent with a complex karyotype. In the second case, WGS detected a complex karyotype comprising one trisomy and two non-classifying balanced translocations, whereas SoC detected the trisomy and only one of the translocations. Since complex karyotypes frequently contain ambiguities, such as marker chromosomes, a direct comparison of specific aberrations between SoC and WGS is futile in cases with three or more SVs/CNVs, independent of the presence of classifying gene-rearrangements. We therefore continued the comparison with the 210 patients who had fewer than three large CNVs and/or SVs according to SoC. Within this cohort, 83/92 (90%) non-classifying SVs/CNVs detected by SoC were also found by WGS (Figure 2). Of eight CNVs missed by WGS, all were present in only 10-25% of the metaphases analyzed with CBA. The remaining undetected aberration was a balanced translocation t(1;2)(p3?4;p2?2) identified by CBA in all metaphases but not in paired WGS analysis. The translocation was detected in unpaired tumor-only re-analysis suggesting a germline origin. WGS detected two SVs and seven CNVs not identified by SoC. One of the SVs was likely cryptic in CBA, while the other SV and 5/7 CNVs showed evidence of low frequency. In one patient, WGS demonstrated loss of chromosome 7. However, SoC revealed an underlying tetraploid karyotype with 4–5 copies of most chromosomes and only 3 copies of chromosome 7, a ploidy pattern not fully captured by WGS. In the last case with excess material corresponding to 7p detected by WGS, CBA was not successful in SoC.

**Figure 2.**
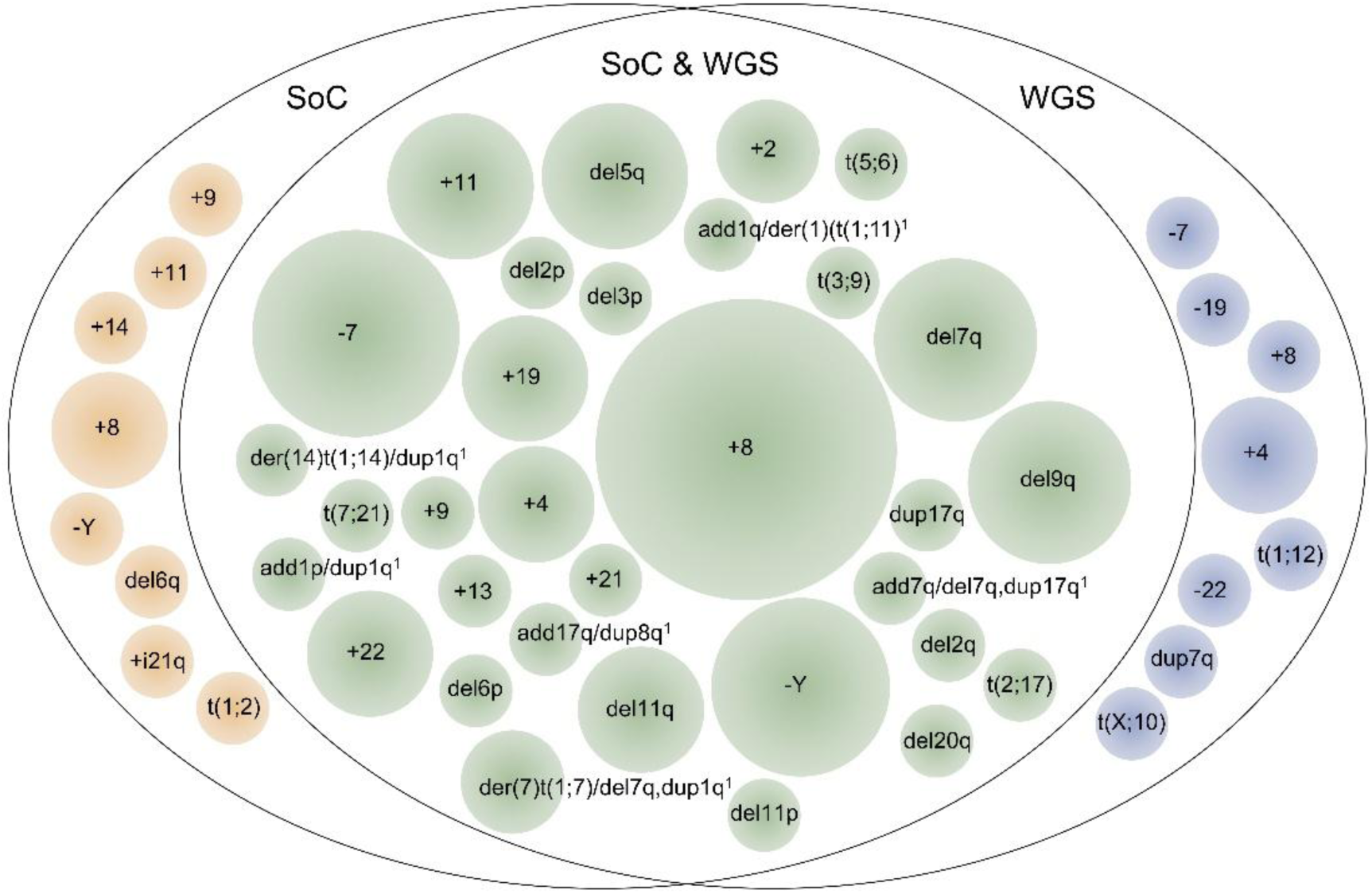
Detection of CNVs and non-classifying structural variants by SoC and by WGS. Circle area in proportion to the number of cases. ^1^The aberration was detected with some differences in SoC compared to WGS but were considered as equivalent findings.

Taken together, WGS is generally well suited for the detection of other SVs and CNVs but poses challenges in situations with subclonality, low tumor content and/or tetraploidy.

### Accurate detection of clinically relevant gene mutations with WGS except for *FLT3*-ITDs at low allele frequency

To assess the detection capacity for SNVs/indels relevant for diagnostic classification, targeted therapy and/or risk-stratification, WGS was compared with SoC targeted PCR or NGS panels. SoC analyses identified 371 SNVs or indels (Figure 1 and Supplementary Table 4), of which 348 (94%) were detected by WGS. Of note, all *NPM1* (*n* = 69), *CEBPA* bZIP-inframe (*n* = 9), and *IDH1/2*-mutations (*n* = 46) were identified by WGS. *FLT3-*ITDs were detected with high confidence by WGS when present at a moderate to high allele frequency. All *FLT3*-ITDs with AR >0.1 were detected, but 16 *FLT3*-ITDs (AR median 0.025, range, 0.01–0.08), were missed. When all *FLT3*-ITDs identified by SoC were considered, WGS correctly classified positive *FLT3*-ITD mutational status in 58/67 (87%) patients (Figure 1), and when applying the AR cutoff of ≥0.05 in 58/61 (95%) patients.

In addition, five mutations in other genes were not detected by WGS, likely due to low VAFs: one *CEBPA*^other^ mutation (VAF 6%), two *TP53* mutations (VAF 5% and 7%), one *BCOR* mutation (VAF 5%), and one *FLT3* mutation in the tyrosine kinase domain (*FLT3*-TKD, VAF 6%). Furthermore, in one patient two presumably clonal hematopoiesis mutations (*BCOR* and *SRSF2)* were missed in the paired WGS analysis due to their presence in the T-cells used as matched germline material. WGS also detected three mutations not reported by SoC: a *STAG2* deletion spanning intronic and exonic regions, an indel in *RUNX1* and a *FLT3*-TKD mutation (VAF 5%). All three were verified upon inspection of SoC raw data.

### Unpaired WGS analysis is sufficient in most cases

In this study, the majority (74%) of WGS analyses were performed in a paired ‘tumor-normal’ setting, with ‘normal’ material from peripheral T cells, cultured fibroblasts, or remission samples. While a paired analysis is required for genome-wide somatic characterization, we aimed to determine whether a tumor-only approach is sufficient to detect all diagnostic, prognostic and treatment-stratifying variants. The analysis was based on the 60 tumor-only cases and, as in previous analyses, non-classifying SVs and CNVs were evaluated only in cases without complex karyotype (all cases with a complex karyotype identified by SoC also showed profiles interpreted as complex based on WGS). In this subcohort, WGTS identified 132/143 (92%) variants detected by SoC (Supplementary table 5). Discrepant findings included the aforementioned *KMT2A* rearrangement, two CNVs at low allele frequency and eight *FLT3*-ITDs (AR ≤0.08) not detected by WGTS, as well as two AML-classifying gene rearrangements (*DEK*::*NUP214* and *PICALM*::*MLLT10*) and three trisomies detected only by WGTS. This result indicates that tumor-only analysis performs similarly to paired analysis for detection of diagnostic, prognostic and treatment stratifying variants.

### Ring trial for data analysis and interpretation

This study was performed in a decentralized setting where WGTS, bioinformatic analysis, interpretation and reporting were performed at four diagnostic centers. The high concordance in genetic variants detected by WGTS compared to SoC indicates comparable performance at all sites. To further benchmark analysis pipelines and data handling across sites, data from seven cases, analyzed by paired tumor-normal WGS and tumor WTS, were shared with the four sites for blinded data analysis and interpretation. The cases comprised gene rearrangements, CNVs, SNVs and indels, and included more challenging aberrations such as a large *FLT3*-ITD and a *MECOM*-rearrangement (Supplementary table 6). For all seven cases, the clinically relevant alterations were reported from all sites. Minor differences in reports included annotation of the *FLT3*-ITD and of SVs (i.e. the *MECOM*-rearrangement as inv(3;3) vs. t(3;3) and a *KMT2A*::*MLLT10* as ins(11;10) vs. t(10;11)). These results demonstrate consistent performance across all four centers in detecting and interpreting clinically relevant genetic alterations in patients with AML.

### Increase in genetically defined AML entities with WGTS

Since one of the primary purposes of genetic analyses is to enable accurate diagnostic classification, we translated the genetic findings from WGTS and SoC into AML subclasses according to WHO 5^th^ and ICC. Using the WHO classification, 222 cases (94%) were identically classified by WGTS and SoC, and 216 cases (92%) using ICC (Figure 3). Most of the observed discrepancies were due to gene rearrangements detected by WGTS but missed by SoC: 10 cases (4%) according to WHO 5^th^ and 12 cases (5%) according to ICC (Supplementary table 7). Additional discordant cases were described in previous sections and included the patient with presumably false positive *KMT2A* rearrangement by SoC, gene mutations missed by paired WGS, erroneous monosomy 7 and two cases with presumably subclonal +8 detected either only by WGS or only by SoC. In summary, WGTS improved genetic subclassification in 5–6% (WHO 5^th^, *n* = 11, ICC, *n* = 14) of patients, while failed to detect clinically relevant findings in only 1% (WHO 5^th^, *n* = 2 WHO, ICC, *n* = 2).

**Figure 3.**
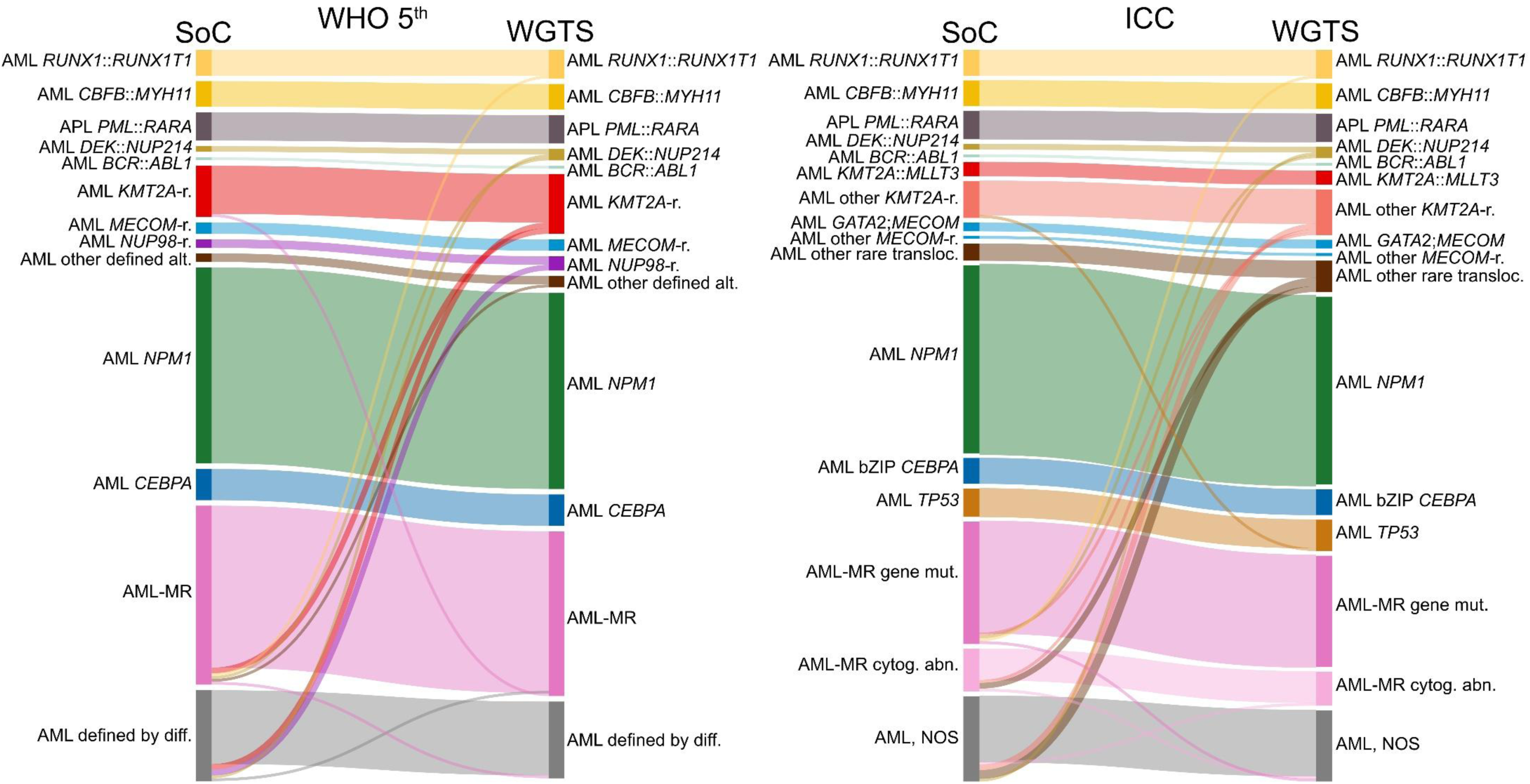
AML classification by SoC and by WGTS. Sankey chart comparing diagnostic classification by WHO 5^th^ edition and ICC. Flow line colors represent the entity interpreted as correct. Diagram created using SankeyMATIC.

### Risk stratification is improved by WGTS

To evaluate the utility of WGTS for genetic risk assessment, all AML patients excluding cases of acute promyelocytic leukemia, APL, independent of age (*n* = 225), were risk stratified according to ELN criteria^3^. Based on results from SoC, 88 patients (39%) were assigned to adverse risk, 78 (35%) to intermediate, and 59 (26%) to low risk (Figure 4). Based on the WGTS results, risk stratification was concordant in 218 (97%) of patients. WGTS improved stratification in 5 patients (2%) attributable to detection of recurrent gene rearrangements. Three patients moved from intermediate to adverse risk due to newly detected gene rearrangements (two *KMT2A* and one *DEK*::*NUP214* rearrangement, Supplementary Table 7). One patient shifted from adverse to favorable risk following identification of a cytogenetically cryptic *RUNX1*::*RUNX1T1* rearrangement, while another shifted from adverse (complex karyotype by SoC) to intermediate risk based on detection of *RUNX1*::*CBFA2T3* by WGTS. One high-risk case was incorrectly reclassified as intermediate risk based on WGTS due to the failure to detect *BCOR* and *SRSF2* mutations (present in the paired normal sample T-cells). In addition, the missed tetraploidy led to incorrect interpretation of chromsosome 7 loss by WGTS, resulting in incorrect stratification of this intermediate case as high-risk. For the pediatric cases (*n* = 38), risk stratification was also performed according to the pediatric trial protocol CHIP-AML22^17^ which resulted in complete concordance between SoC and WGTS. As described above, WGTS missed several low-frequency *FLT3*-ITDs, although this did not influence risk stratification according to ELN or CHIP-AML22. When all *FLT3*-ITDs were considered (lowering the AR cutoff from 0.05 to 0.01 in SoC^18^), three cases would be reclassified based on missed *FT3*-ITD by WGTS: two adults from ELN intermediate to favorable risk (Supplementary figure 1) and one child from CHIP-AML22 high-risk to standard-risk. The remaining patients with missed *FLT3*-ITD (all adults) were risk-stratified based on other genetic aberrations (Figure 1). In conclusion, WGTS performs well for risk stratification in AML.

**Figure 4.**
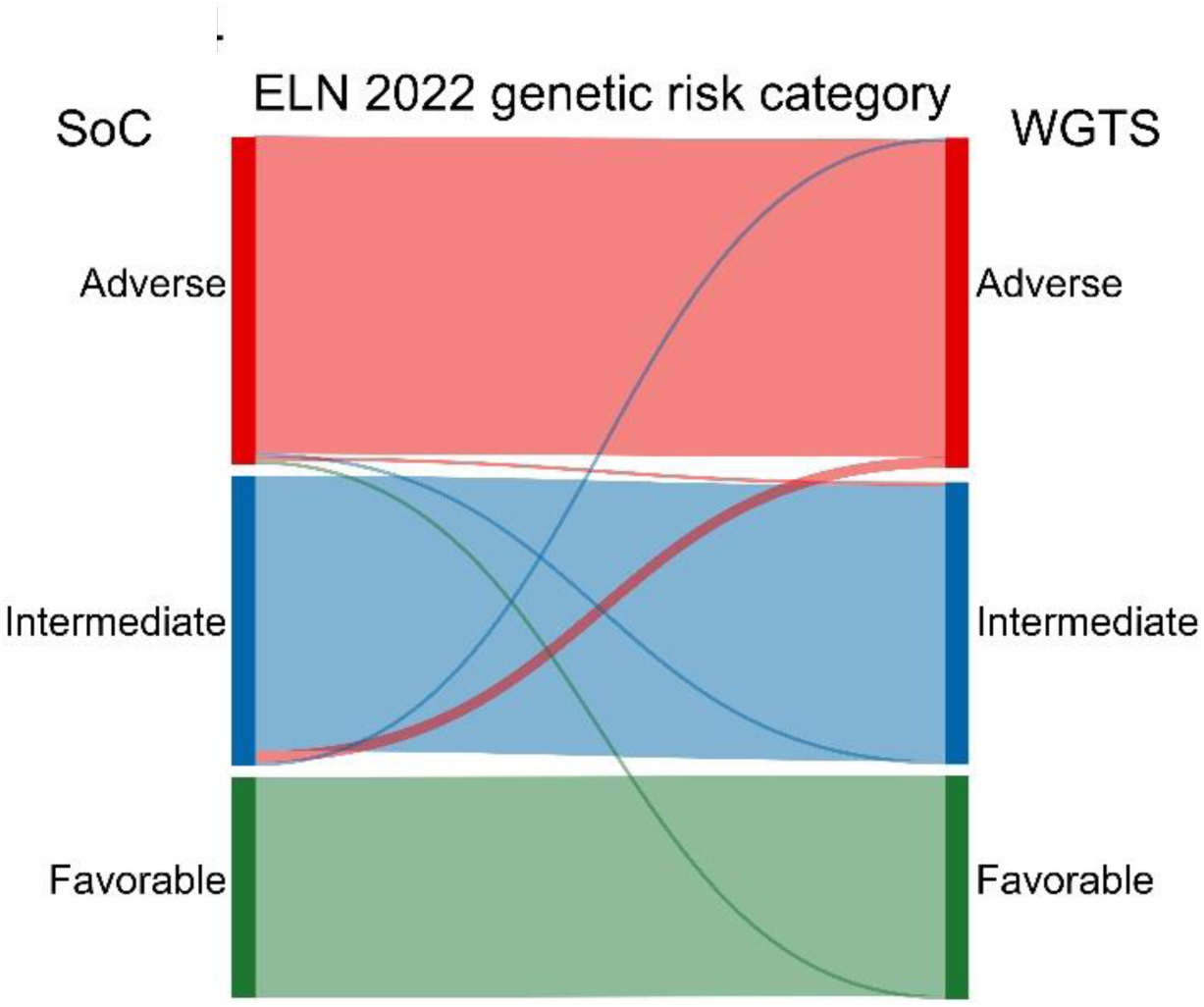
ELN 2022 risk classification by genetics by SoC and by WGTS. Includes all AML patients (*n* = 225) excluding acute promyelocyte leukemia, APL. *FLT3*-ITD AR≥0.05 used for stratification. Flow line colors represent the risk category interpreted as correct. Diagram created using SankeyMATIC.

## Discussion

Untargeted genomic approaches such as WGS and WTS are becoming increasingly accessible and less costly, positioning them as attractive techniques for clinical AML diagnostics. We here show that WGTS captures most genetic aberrations, provides additional clinically relevant information as compared with current SoC diagnostics in 5-6% of cases, and can be implemented in a decentralized manner.

In this study, WGTS identified all but one of the diagnostic and risk-stratifying rearrangements detected by SoC and identified the last SoC-detected rearrangement as a false positive result, corresponding to a sensitivity of 100%. In addition, WGTS revealed 12 additional rearrangements, increasing the detection of diagnostically relevant rearrangements in 6% (13/235) of cases, including the identification of *NUP98* and *KMT2A* rearrangements. Although not included in ELN2022, recent data identifies *NUP98* rearrangements as associated with a high risk of relapse^19^ and they are both of high relevance for targeted treatment with menin inhibitors^20^. The improved results of WGTS were driven by the high resolution of sequencing, enabling detection of cytogenetically cryptic rearrangements as well as precise characterization of the underlying aberrations responsible for abnormal FISH signals. Increased detection of gene fusions compared with CBA has also been reported by Voss et al who compared an integrated WGTS approach with cytogenetics in pediatric AML patients, and by Duncavage et al, who compared WGS with cytogenetics and FISH in adult AML and MDS patients^4,9^. Although this study used WGTS, all gene-rearrangements identified by WTS were also detected by WGS. In addition, WTS missed several rearrangements detected by WGS, including *MECOM* rearrangements, as previously reported by Voss et al^9^, and t(7;12). Both these entities are characterized by dysregulated gene expression rather than a fusion gene, driven by enhancer hijacking in AML with *MECOM* rearrangement^21,22^, and by *MNX1* overexpression in AML with t(7;12)^23^. Despite its limited added value upfront, WTS remains a useful complement to WGS for validating rearrangements and reduces the need for targeted PCR assays. Additionally, an integrated WGS/WTS variant calling may further improve sensitivity^9,12^.

For other SVs and large CNVs, WGS had a sensitivity of 90%, which is lower than that observed for gene fusions. Complex karyotypes were generally well captured by WGS and most discrepancies when compared to SoC consisted of additional or corrected gene rearrangements. Only one case reached the definition of a complex karyotype solely by WGS, this comparison is, however, hampered by the lack of an established WGS definition for complex karyotype. Variants (CNVs) missed by WGS or SoC were predominantly subclonal. This can be expected due to selective growth advantages of clones *in vitro* during culturing preceding CBA. Even though WGS remains challenging in rare cases with tetraploidy, its capacity to resolve complex rearrangements is of greater importance, particularly as it can uncover aberrations with diagnostic or risk-stratifying implications.

In our study, WGS achieved a sensitivity of 94% for gene mutations, including both SNVs and indels. The most clinically relevant aberrations missed were *FLT*3-ITDs, primarily those with low allele frequency. This aligns with findings by Voss et al, who found that most *FLT*3-ITDs with AR below 0.1 were either undetected or identified with low certainty by WGS alone; high sensitivity – comparable to PCR - was only achieved by combining WGS with WTS^9^. Reliable detection of variants with low VAF would require an increased sequencing depth at a substantially higher cost, currently limiting its feasibility for clinical diagnostics. In addition, turnaround time requirements for initiating targeted therapy with FLT3 inhibitors cannot be met in our clinical WGTS setting. Thus, a parallel high-sensitivity targeted assay for *FLT3*-ITD, such as PCR as used in previous WGS-based decision-making algorithms^4^, remains necessary. Another cause of missed variants by WGS was the paired tumor–normal analysis, which obscured relevant mutations such as clonal hematopoiesis associated variants in *BCOR* and *SRSF2*. This is consistent with reports of pre-leukemic events being present in lymphoid cells^24,25^ and underscores the limitations of using T cells as normal control. The paired analysis pipeline also failed to detect a germline nonrecurrent constitutional translocation. Notably, all these variants were identifiable with tumor-only analysis, as demonstrated in the subset of patients lacking matched normal material. In a clinical setting, we therefore favor an initial, rapid tumor-only analysis, complemented by a comprehensive paired analysis using DNA from cultured fibroblasts.

The blinded ring trial for WGTS data analysis showed a high degree (100%) of concordance between sites, with only minor discrepancies in annotation. Although none of the discrepancies would have affected patient management, they highlight opportunities to further optimize bioinformatic pipelines and reduce reliance on manual interpretation. Overall, these findings demonstrate the feasibility of a decentralized WGTS approach for consistent detection of clinically relevant genetic alterations.

Diagnostic classifications according to WHO 5^th^ and ICC were very similar between WGTS and SoC, although a more accurate classification was achieved in 5–6% of patients due to WGTS-identified rearrangements. ELN risk group assignment was also highly concordant, with WGTS leading to re-stratification in only 2% of cases. The few discrepancies in which WGTS missed risk-stratifying aberrations were attributable to the paired tumor–normal workflow, type of normal material, and challenges in distinguishing numerical abnormalities from tetraploidy. For comparison, the adult ELN risk classification was applied to all AML patients using an AR cutoff of 0.05 for *FLT3*-ITD^26^. Using this approach, WGTS did not alter *FLT3*-ITD based stratification. The benefit of WGTS for risk classification may be even greater in pediatric AML where treatment protocols differ but commonly include *FLT3*-ITD and a varying number of fusion genes for guidance of treatment intensity^27^. In this study with a limited number of children, WGTS did not alter identification of risk-adapting genetic aberrations according to the CHIP-AML22 treatment protocol.

In conclusion, WGTS improves the detection of clinically relevant genetic alterations compared with SoC and can be implemented at a national scale in a decentralized setting, facilitating logistics while strengthening local expertise and communication between clinicians and diagnostic laboratories. The costs associated with our workflow^28^ and a turn-around time of 10– 14 working days, as reported by others^29,30^, appears feasible and clinically acceptable when complemented by specific targeted assays to support for early treatment decisions. In Sweden, WGTS is since 2024 implemented for all children with cancer, including acute leukemias^31^. Implementation for adult acute leukemia is currently being performed as a real-time validation study, evaluating turn-around times and health economics, with the ultimate goal to replace most current SoC assays.

## Data Availability

Access to patient-level genomic data is subject to applicable ethical and data-protection regulations. Genomic data is available from the corresponding author upon reasonable request and appropriate institutional approval.

## Acknowledgements

We thank the staff at Clinical Genetics, Umeå University Hospital, Clinical Genetics and Genomics, Sahlgrenska University Hospital, Genomic Medicine Center Karolinska, Karolinska University Hospital, Section of Molecular Diagnostics, Skåne University Hospital, Molecular Diagnostics and Clinical Genetics, Uppsala University Hospital, and SciLifeLab Clinical Genomics. The study was supported by reagents from Illumina, and grants from the Swedish Childhood Cancer Fund, The Ministry of Health and Social Affairs and Västra Götalandsregionen, the agreement between the Swedish government and the county councils; the ALF-agreement (ALFGBG-1006375).

## Authorship Contributions

LC, LP, VW, GB, TF, LF, and RR planned the study. VL, MJä, SD, AR, MH, GO, JA, CJP, JP, HV, KBS, and UNN participated in patient inclusion. AS, LC, KO, COP, TP, IG, LOA, LS, AK, SP, PB, GB, TF, and LF contributed SoC. AS, LC, COP, TP, LOA, AK, SP, NH, KM, MJo, PB, GB, TF and LF contributed WGTS data. AS, KO and LF performed central review and data analysis. AS, LC, KO, LP, LF, and RR wrote the manuscript. All authors read and approved of the manuscript.

