## Supplementary material for "Clinical validation of whole-genome and transcriptome sequencing for genetic diagnostics in adult and pediatric AML"

### Supplementary tables

**Supplementary table 1.** Sample material for the 235 included AML cases.

| | | Number of patients | Percentage of patients |
|---|---|---|---|
| Tumor material WGS | Bone marrow | 231 | 98% |
| | Peripheral blood | 4 | 2% |
| Normal material WGS | Skin fibroblasts | 74 | 31% |
| | Sorted T-cells | 56 | 24% |
| | Remission sample | 45 | 19% |
| | Not available | 60 | 26% |
| Tumor material WTS | Bone marrow | 189 | 80% |
| | Peripheral blood | 4 | 2% |
| | Not available | 42 | 18% |

**Supplementary table 2.** Bioinformatic tools and interpretation software used.

| | **Stockholm** | **Uppsala** | **Lund** | **Gothenburg** |
|---|---|---|---|---|
| Workflow definition framework | Snakemake | Snakemake | NextFlow | Snakemake |
| Reference genome | GRCh37 (hg19) | GRCh38 (hg38) | GRCh38 (hg38) | GRCh38 (hg38) |
| WGS Aligner | Sentieon bwa-mem | Parabricks bwa-mem | Sentieon bwa-mem | Sentieon bwa-mem |
| WGS SNV/indel caller | Sentieon-TNScope | Parabricks mutectcaller, Pindel | Vardict, Freebayes | Sentieon-TNScope, Pindel |
| WGS CNV caller (visual inspection IGV or similar tools) | AscatNGS | CNVKit, GATK's somatic CNV detection pipeline | GATK's somatic CNV detection pipeline | Canvas |
| WGS SV caller | Manta, Delly, Tiddit | Manta | Manta | Manta |
| WTS aligner | STAR | STAR | STAR | STAR |
| WTS fusion gene caller | STARfusion, FusionCatcher | STARfusion, FusionCatcher, Arriba | STARfusion, FusionCatcher, Arriba | STARfusion, FusionCatcher, Arriba |
| Tool for clinical interpretation | Scout[1] | QCI (Qiagen) | Coyote[1] | QCI (Qiagen) |
| Code availability | https://github.com/Clinical-Genomics/BALSAMIC https://github.com/Clinical-Genomics/scout https://github.com/nf-core/rnafusion | https://github.com/hydra-genetics/ https://github.com/clinical-genomics-uppsala/fluffy_hematology_wgs | https://github.com/SMD-Bioinformatics-Lund/nextflow_tumwgs https://github.com/SMD-Bioinformatics-Lund/coyote https://github.com/SMD-Bioinformatics-Lund/nextflow_rnaseqfus | https://github.com/ClinicalGenomicsGBG/wgs_somatic https://github.com/ClinicalGenomicsGBG/qd_rna |

[1] In-house developed software.

**Supplementary table 3**. Mandatory genetic aberrations investigated in the study.

| **Cytogenetic aberrations** | | | **Gene mutations** | |
|---|---|---|---|---|
| Complex karyotype | del(12p)/t(12p)/add(12p) | *KMT2A*-rear. | *ASXL1* | *NPM1* |
| -5, del(5q), t(5q) | idic(X)(q13) | *MECOM*-rear. | *BCOR* | *RUNX1* |
| -7 or del(7q) | Any CNV > 5 Mb | *DEK*::*NUP214* | *CEBPA* | *SF3B1* |
| -17/abn(17p) | *PML*::*RARA* | *RBM15*::*MRTFA* | *EZH2* | *SRSF2* |
| del(11q) | *RUNX1*::*RUNX1T1* | *BCR*::*ABL1* | *FLT3* | *STAG2* |
| del(20q) | *CBFB*::*MYH11* | Other classifying rearrangements* | *IDH1* | *TP53* |
| -13/del(13q) | *NUP98*-rear. | | *IDH2* | *U2AF1* |

***RUNX1T3*(*CBFA2T3*)::*GLIS2*/inv(16)(p13q24, *FUS*::*ERG*/t(16;21)(p11;q22), *MNX1*::*ETV6*/t(7;12)(q36;p13), *NPM1*::*MLF1*/t(3;5)(q25;q35), *PRDM16*::*RPN1*/t(1;3)(p36.3;q21.3), *PICALM*::*MLLT10*/t(10;11)(p12.3;q14.2) or *RUNX1*::*CBFA2T3*/t(16;21)(q24.3;q22.1).

**Supplementary table 4**. Gene mutations as detected by targeted PCR and NGS-panel in standard of care (SoC) and by WGS.

| **SoC only** | **SoC and WGS** | | **WGS only** |
|---|---|---|---|
| *BCOR* | *NPM1* (*n* = 69) | *ASXL1* (*n* = 19) | *STAG2* |
| *SRSF2* | *CEBPA*-bZIP$^{\text{inframe}}$ (*n* = 9) | *BCOR* (*n* = 11) | *RUNX1* |
| *CEBPA*$^{\text{other}}$ | *CEBPA*$^{\text{other}}$ (*n* = 22) | *RUNX1* (*n* = 28) | *FLT3*-TKD |
| *TP53* (*n* = 2) | *TP53* (*n* = 16) | *SF3B1* (*n* = 3) | |
| *BCOR* | | | |
| *FLT3*-TKD | *IDH1/2* (*n* = 46) | *SRSF2* (*n* = 14) | |
| *FLT3*-ITD (*n* = 16) | *FLT3*-ITD (*n* = 65) | *STAG2* (*n* = 13) | |
| | *FLT3*-TKD (*n* = 22) | *U2AF1* (*n* = 8) | |
| | | *EZH2* (*n* = 3) | |

**Supplementary table 5**. Detection of variants by SoC and WGTS in the subcohort of 60 cases with analyses performed as tumor only.

| **SoC only** | **SoC and WGTS** | | **WGTS only** |
|---|---|---|---|
| *KMT2A*-rear. | *KMT2A*-rearr (n = 2) | *NUP98*-rearr | *DEK*::*NUP214* |
| | *MECOM*-rear (n = 2) | *RUNX1*::*RUNX1T1* | *PICALM*::*MLLT10* |
| | *CBFB*::*MYH11* | *PML*::*RARA* (n = 5) | |
| +8 | +4 | del(5q) (n = 3) | -7 |
| del(6q) | -7 (n = 4) | del(7q) (n = 2) | -19 |
| | +8 (n = 3) | del(9q) | -22 |
| | +11 | del(11q) | |
| | +22 | del(20q) | |
| | t(7;21) | der(14)t(1;14) / dup(1q) [1] | |
| *FLT3*-ITD | *NPM1* (n = 11) | *ASXL1* (n = 7) | |
| (n = 8) | *CEBPA*- bZIP$^{\text{inframe}}$ (n = 3) | *BCOR* (n = 6) | |
| | *CEBPA*$^{\text{other}}$ (n =2) | *RUNX1* (n = 13) | |
| | *TP53* (n = 9) | *SF3B1* | |
| | *IDH1/2* (n = 9) | *SRSF2* (n = 6) | |
| | *FLT3*-ITD (n = 20) | *STAG2* (n = 6) | |
| | *FLT3*-TKD (n = 5) | *EZH2* (n = 2) | |

[1]The aberration was detected with some differences in SoC compared to WGS but were considered as equivalent findings.

**Supplementary table 6**. Results from ring trial for WGTS data analysis and interpretation. Fastq-files (WGS tumor, WGS normal and WTS tumor) were shared to the four sites.

| Case | Aberrations | Site 1 | Site 2 | Site 3 | Site 4 |
|---|---|---|---|---|---|
| #1 | *NPM1*:c.860_863dup p.(Trp288Cysfs*12) | √ | √ | √ | √ |
| | *FLT3*:c.1770_1793dup p.(Tyr597_Glu598ins8) | √ | √ | √ | √ |
| | *FLT3*:c.2028C>G p.(Asn676Lys) | √ | √ | √ | √ |
| | *FLT3*:c.2503G>C p.(Asp835His) | √ | √ | √ | √ |
| | *IDH2*:c.419G>A p.(Arg140Gln) | √ | √ | √ | √ |
| #2 | del(11)(q14.1) | √ | √ | √ | √ |
| | del(Y)(q11.1) | √ | √ | √ | √ |
| | *FLT3*:c.1906_1907_ins261 p.(Thr635_Gly636ins87) | √ | √ | √[1] | √ |
| | *RUNX1*:c.497_508+1dup p.(Ser172Trpfs*45) | √ | √ | √ | √ |
| | *RUNX1*:c.408T>G p.(Asn136Lys) | √ | √ | √ | √ |
| | *SRSF2*:c.284_307del p.(Pro95_Arg102del) | √ | √ | √ | √ |
| #3 | *KMT2A*::*MLLT3*, t(9;11)(p21.3;q23.3) | √ | √ | √ | √ |
| #4 | dup(1)(q10q44) | √ | √ | √ | √ |
| | del(7)(q10) | √ | √ | √ | √ |
| | *IDH2*:c.515G>A p.(Arg172Lys) | √ | √ | √ | √ |
| #5 | *KMT2A*::*MLLT10*, der(11)ins(11;10)(q23.3;p12.31p12.31) | √[2] | √[2] | √[2] | √ |
| | *ASXL1*:c.1934dup p.(Gly646Trpfs*12) | √ | √ | √ | √ |
| | *U2AF1*:c.470A>C p.(Gln157Pro) | √ | √ | √ | √ |
| #6 | *GATA2*::*MECOM*, inv(3)(q21.3;q26.2) | √ | √[3] | √ | √ |
| | -7 | √ | √ | √ | √ |
| | *BCOR*:c.1282G>T p.(Glu428*) | √ | √ | √ | √ |
| #7 | *RUNX1*::*RUNX1T1*, t(8;21)(q21.3;q22.12) | √ | √ | √ | √ |
| | del(9)(q21.11q21.33) | √ | √ | √ | √ |

[1] Detected with alternate annotation

[2] Detected as t(10;11)(p12.31;q23.3)

[3] Detected as t(3;3)(q21.3;q26.2)

**Supplementary table 7**. Genetic findings by SoC and by WGTS for cases where discrepant findings resulted in altered diagnostic classification or altered risk stratification. Adult cases shown as purple, children as yellow. Discrepant findings altering diagnosis and/or risk stratification highlighted in bold.

| No. | Genetics by SoC (Gene mutations annotated on MANE select transcript) | Classification by SoC (WHO) | Classification by SoC (ICC) | ELN risk | Genetics by WGTS (Gene mutations annotated on MANE select transcript) | Classification by WGTS (WHO) | Classification by WGTS (ICC) | ELN risk |
|---|---|---|---|---|---|---|---|---|
| #1 | add(X)(p22),t(8;16)(p11;p13)/47,idem,+19/48, idem,+19,+21 | AML MR | AML MR cytogenetics | Adverse | t(8;16)(p11;p13),+19,+19,+21, ***KAT6A::CREBBP*** | AML other def. genetic alt. | AML other rare recur. translocations | Adverse |
| #2 | -X,add(6)(p25),-11,+mar1,+mar2, +mar3[6]/47,idem,-12,+mar1[2]/ 46,idem,-12, add(13)(q32),-14,+mar2 | AML MR | AML MR cytogenetics | Adverse | t(X;11;10)(q24;q23.3:p12.31), dup(1)(q21.1q44),dup(5)(p12pter) ,del(12)(p12.3), ***KMT2A::MLLT10*** | AML *KMT2A*-r | AML other *KMT2A*-r | Adverse |
| #3 | i(21)(q10), *ASXL1*:c.1934dup, p.(G646Wfs*12) *U2AF1*:c.470A>C, p.(Q157P) | AML MR | AML MR gene mut. | Adverse | t(10;11)(p12.31;q23.3) ***KMT2A::MLLT10*** *ASXL1*:c.1934dup, p.(G646Wfs*12) *U2AF1*:c.470A>C, p.(Q157P) | AML *KMT2A*-r | AML other *KMT2A*-r | Adverse |
| #4 | *FLT3*-ITD *U2AF1*: c.467G>A p.(R156H) | AML MR | AML MR gene mut. | Adverse (High-risk*) | t(6;9), ***DEK::NUP214*** *FLT3-ITD* *U2AF1*: c.467G>A p.(R156H) | AML *DEK::NUP214* | AML *DEK::NUP214* | Adverse (High-risk*) |
| #5 | *FLT3*-ITDx2 (total AR > 0,05) | AML, defined by diff. | AML, NOS | Inter-mediate (High risk*) | ***NUP98::NSD1*** *FLT3*-ITD (AR >0,05) | AML *NUP98*-r | AML other rare recur. Translocations | Inter-mediate (High risk*) |
| #6 | *FLT3*-ITD (AR > 0,05) | AML, defined by diff. | AML, NOS | Inter-mediate | ***NUP98::NSD1*** *FLT3*-ITD (AR >0,05) | AML *NUP98*-r | AML other rare recur. Translocations | Inter-mediate |
| #7 | - | AML, defined by diff. | AML, NOS | Inter-mediate | ***KMT2A::ELL*** | AML *KMT2A*-r | AML other *KMT2A*-r | Adverse |
| #8 | - | AML, defined by diff. | AML, NOS | Inter-mediate | ***KMT2A::MLLT10*** | AML *KMT2A*-r | AML other *KMT2A*-r | Adverse |
| #9 | *FLT3*-ITD (AR >0,05) | AML, defined by diff. | AML, NOS | Inter-mediate | ***DEK::NUP214*** *FLT3*-ITD (AR >0,05) | AML *DEK::NUP214* | AML *DEK::NUP214* | Adverse |
| #10 | +8, +13, der(12)t(1;12)(q2?;p13), del(21)(p?) | AML MR | AML MR cytogenetics | Adverse (Standard-risk*) | +8, +13, dup 1q, del 12p, t(16;21)(q24;q22) ***RUNX1::CBFA2T3*** | AML MR | AML other rare recur. Translocations | Inter-mediate (Standard-risk*) |
| #11 | *U2AF1*:c.470A>C p.(Q157P) | AML MR | AML MR gene mut. | Adverse | ***RUNX1::RUNX1T1*** *U2AF1*:c.470A>C p.(Q157P) | AML *RUNX1::RUNX1T1* | AML *RUNX1::RUNX1T1* | Favor-able |
| #12 | add(1)(p36), del(4)(q2?),-5,-7,-11,-13,add(14)(q?),add(19)(p?),-21,+4mar *TP53*:c.733G>A p.(G245S) ***KMT2A*-r** | AML KMT2A-r | AML w. other KMT2A-r | Adverse | del 4q, del 5q, -7, del 14q, del 17p, t(19;21)(q13;q22) t(14;17)(q24p13), t(1;21)(p36q11) *TP53*:c.733G>A, p.(G245S) | AML MR | AML with mut. TP53 | Adverse |

| | | | | | | | | |
|---|---|---|---|---|---|---|---|---|
| #13 | **Tri-tetraploid karyotype** by multiple FISH assays (not conclusive by G-banding) | AML, defined by diff. | AML, NOS | Inter-mediate | +4, -6, -7, +14, -15, -18, +21, +22, dup(17)(q12q25.3), t(9;10;11)(q34.1;p12.31;q14.2), ***PICALM*::*MLLT10*** | AML MR | AML other rare recur. Translocations | Adverse |
| #14 | +14<br>*FLT3*-ITD (AR < 0,05)<br>***BCOR***:c.2811del p.(T938Pfs*18)<br>***SRSF2***:c.284_307del p.(P95_R102del)<br>*IDH2*:c.419G>A p.(R140Q) | AML MR | AML MR gene mut. | Adverse | *IDH2*:c.419G>A p.(R140Q) | AML, defined by diff. | AML, NOS | Inter-mediate |
| #15 | **+8**<br>*FLT3*-ITD (AR > 0,05) | AML, defined by diff. | AML MR cytogenetics | Inter-mediate | *FLT3*-ITD (AR > 0,05) | AML, defined by diff. | AML, NOS | Inter-mediate |
| #16 | *IDH2*:c.515G>A p.(R172K) | AML, defined by diff. | AML, NOS | Inter-mediate | **+8**<br>*IDH2*: c.515G>A p.(R172K) | AML, defined by diff. | AML MR cytogenetics | Inter-mediate |

*grouping based on presence of genetic marker influencing treatment stratification in pediatric treatment protocol CHIP-AML22

#### Supplementary figures

**Supplementary figure 1.** ELN risk stratification of non-APL patients according to SoC vs. WGTS with *FLT3*-ITD AR cutoff at 0.01

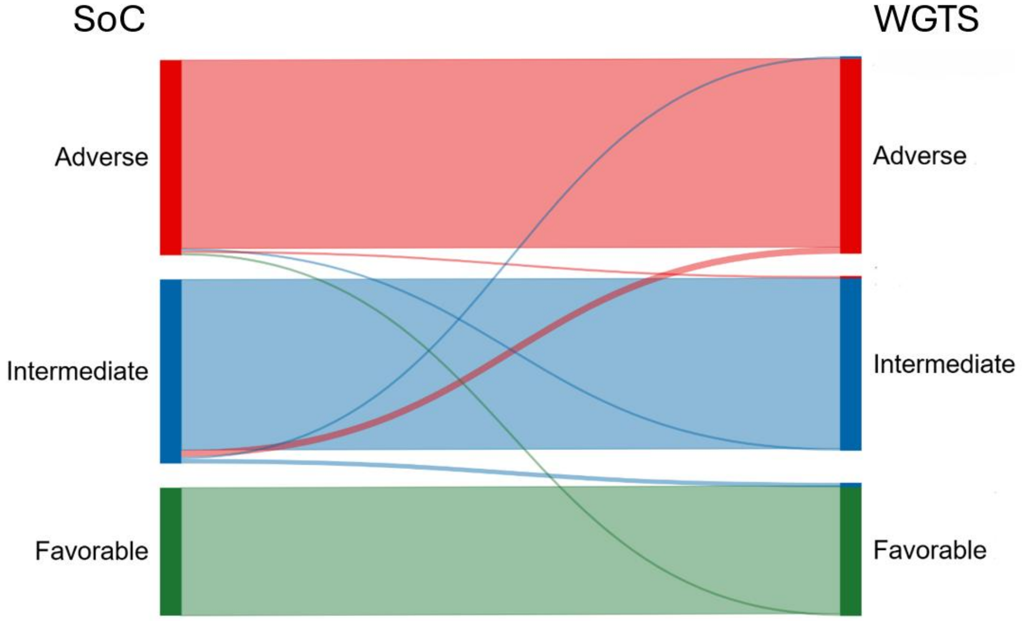
